# Impact of Salivary and Pancreatic Amylase Gene Copy Numbers on Diabetes, Obesity, and Functional Profiles of Microbiome in Northern Japanese Population

**DOI:** 10.1101/2021.10.02.21264452

**Authors:** Takanori Hasegawa, Masanori Kakuta, Rui Yamaguchi, Noriaki Sato, Tatsuya Mikami, Koichi Murashita, Shigeyuki Nakaji, Ken Itoh, Seiya Imoto

## Abstract

Amylase genes reside in a structurally complex locus, and their copy numbers vary greatly, especially among agricultural races. Amylase genes seem to shape the metabolic response to dietary starch, and several studies have reported their association with obesity. Besides, the effect of amylase copy numbers seems to depend on lifestyle, and the mechanism of this effect was partially explained by changes in the oral and gut microbiome compositions; however, a detailed mechanism has been unclarified. In this study, we showed their association with diabetes in addition to obesity, and further discovered a plausible mechanism of this association based on the function of commensal bacterial in a northern Japanese population. First, we confirmed that the amylase copy number in the population tends to be larger than that reported in other studies and that there is a positive association between obesity and diabetes (p =1.95E-2 and 3.28E-2). Second, we identified that relative abundance of some genus level microbiome, *Capnocytophaga, Dialister*, and previously reported bacteria, were significantly associated with amylase copy numbers. Finally, through functional gene-set analysis using shotgun sequencing, we observed that the abundance of genes in the Acarbose pathway in the gut microbiome was significantly decreased with an increase in the amylase copy number (p-value = 5.80E-4), which can partly explain the mechanism underlying obesity and diabetes in populations with high amylase copy numbers.

## Introduction

Similar to hundreds of human genes, the amylase genes reside in a structurally complex locus, with inversions, deletions, and many duplications. Copy number variation (CNV) of AMY genes, which encode amylase enzymes that digest starch into maltose and dextrin, is considered one of the strongest signals of recent natural selection in the human population. There are three amylase genes; the copy number of AMY1 (AMY1-CN), which encodes salivary amylase, varies from 1 to more than 20 copies, whereas the copy numbers of AMY2A (AMY2A-CN) and AMY2B (AMY2B-CN), which encode pancreatic amylase, vary from 1 to more than 6 copies. AMY1-CN is known to be positively correlated with salivary amylase activity and hydrolyzes the alpha bonds of starch and glycogen, initiating the process of starch degradation in the mouth. As the variety in the number of these genes is specific to agricultural races among humans, the CNVs of these three genes are hypothesized to shape the metabolic response to dietary starch (1-3).

In recent decades, several studies have shown that AMY1-CN and salivary amylase concentrations are associated with obesity and diabetes. Further, an increase in AMY1-CN levels is reported to decrease the risk of obesity (4-9). However, some other studies have reported conflicting results and showed no or contrasting associations of AMY1-CN with obesity (10-12). These conflicting results may be due to the heterogeneity in patient samples in terms of genetic background and lifestyle, as well as the use of different study methods, such as the assessment of AMY1-CN. Interestingly, Rukh et al. reported that the degree of dietary starch intake modifies the relationship between copy number variation in the salivary amylase gene and BMI; thus, an increase in AMY1-CN decreases the risk of obesity in the low starch intake group and increases the risk in the high starch intake group (12). Marcovecchio et al. also showed that an increase in AMY1-CN decreases the risk of obesity in boys, but can increase its risk in girls (9). Some other studies have shown that even though AMY1-CN is associated with salivary amylase levels, the salivary amylase level is more associated with obesity and BMI compared to AMY1-CN (13-15).

Humans with high AMY1-CN (AMY1H), who can produce high levels of salivary amylase, might thus derive more energy from the same amount of diet compared to those with low AMY1-CN (AMY1L). Compared to AMY1H, AMY1L might be expected to harbor gut microbiomes with a greater capacity for the decomposition of carbohydrates. Thus, AMY1 copy number has also recently emerged as a significant factor in shaping the relative abundance of the specific members in the human oral and gut microbiota (16-18). Further, an experimental study elucidated that the types of microbes in AMY1H drive higher adiposity when transferred to germ-free mice (18), but the detailed mechanism has not been clarified.

In this study, we aimed to investigate the fundamental relationships of AMY1 and AMY2A with diabetes and obesity in a homogenous northern Japanese population considered to have a high starch intake. We first identified AMY1-CN, AMY2A-CN, and AMY2B-CN, and their mutation distributions using whole-genome sequencing data. In many studies, CNV arrays have been used to detect CNVs; however, recent studies suggest that whole-genome sequencing (WGS) analysis of CNVs is likely to be more efficient and outperforms array-based CNV analysis (19). We utilized a methodology that has been developed to accurately estimate CNVs in AMY regions from sequencing data (20, 21). Secondly, we analyzed the association of the copy numbers of AMY genes (AMY-CNs) with obesity and diabetes. We then analyzed the oral and gut microbiome at the genus level to discriminate high and low AMY-CN individuals using 16S rRNA gene sequence analysis. Finally, we obtained shotgun whole-genome sequencing data of the oral and gut microbiome to investigate the functional gene sets of starch degradation pathways that can be related to AMY-CNs.

## Results

### Distribution of AMY1, AMY2A, and AMY2B copy number variation

We first analyzed the WGS data to detect AMY-CN from 1,479 individuals in the 2015 and 2016 participants. For estimating AMY-CN, we applied TIDDIT (20) and AMYCNE (21), and then obtained AMY1A, AMY2A, and AMY2B CNs and their relationships, as illustrated in Fig. 1. In contrast to the average AMY1-CN 10.3 in this population, as described in Fig. 1(a), previous studies have reported much smaller average AMY1-CNs; for example, the average CN was 7 in a study from Qatar (13), 8.3 in a study from Japan (14), 7.9 in a study from the US (18), and 7.0 in a study from Mexico (16). As illustrated in Fig. 1(b), AMY1 and AMY2A CNs were strongly correlated (r = 0.60, p = 2.82E-144) because their genetic regions are near and on the same linkage disequilibrium (LD) block (11). On the contrary, AMY2B-CN was not associated, and almost all participants had two copies. Thus, we focused on AMY1 and AMY2A CNs in the following experiments.

**Figure 1.**
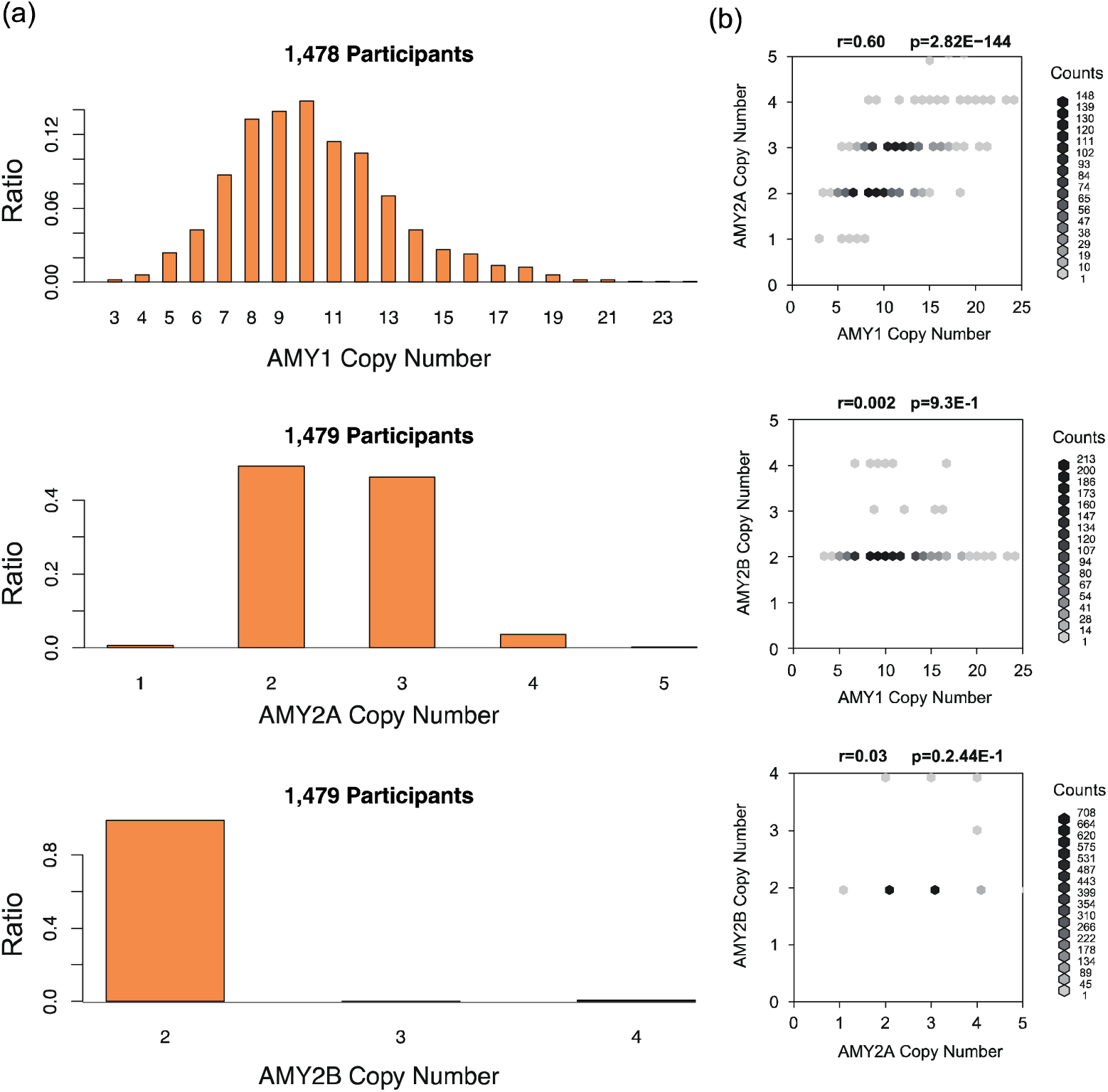
Histogram of AMY1, AMY2A, and AMY2B copy number variations and their associations. In the left of this figure, the histograms of AMY1A, AMY2A, and AMY2B-CNs are displayed. The number of analyzed participants are also indicated at the top of each histogram. In the right of this figure, associations among AMY1A, AMY2A, and AMY2B-CNs are illustrated as a scatter plot. At the top of each scatter plot, *r* and *p* indicate Pearson’s correlation coefficient and p-value, respectively.

### SNP patterns on AMY1, AMY2A and AMY2B regions

Next, we summarized the SNP distributions in the AMY gene regions as shown in Table 1 and illustrated in Fig. 2. Contrary to the results of AMY CNV, the number of SNPs was clearly larger in the AMY2A and AMY2B regions than in the AMY1 region. However, a previous study reported that several SNPs with a minor allele frequency of more than 0.3, for example, rs42443712, were detected in the AMY1A region (11). In addition, several SNPs discovered in our analysis did not have a reference SNP ID (rsID) assigned to an identical location. It could be that the genetic variety of AMY1A might not be observed because this cohort is from a region of Japan where genetic diversity appears to be low. Notably, even though the effects of these SNPs on BMI and HbA1c were investigated, no significant effect was detected, similar to a previous study (11). Thus, we focused on the effects of AMY-CN in the following sections.

**Table 1.** Summary of SNPs detected in the AMY1, AMY2A, and AMY2B gene regions.

|  | exonic (nonsynonymous, synonymous, top) | intronic | UTR5 | splicing | intergenic |
| --- | --- | --- | --- | --- | --- |
| AMY1 | 20 (19, 1, 0) | 1,131 | 0 | 0 | 583,736 |
| AMY2 | 564 (251, 292, 1) | 13,253 | 113 | 1 | 13,456 |
| AMY1-AMY2 | - | - | - | - | 1,229 |
| Total | 564 | 14,384 | 113 | 1 | 609,874 |

**Figure2.**
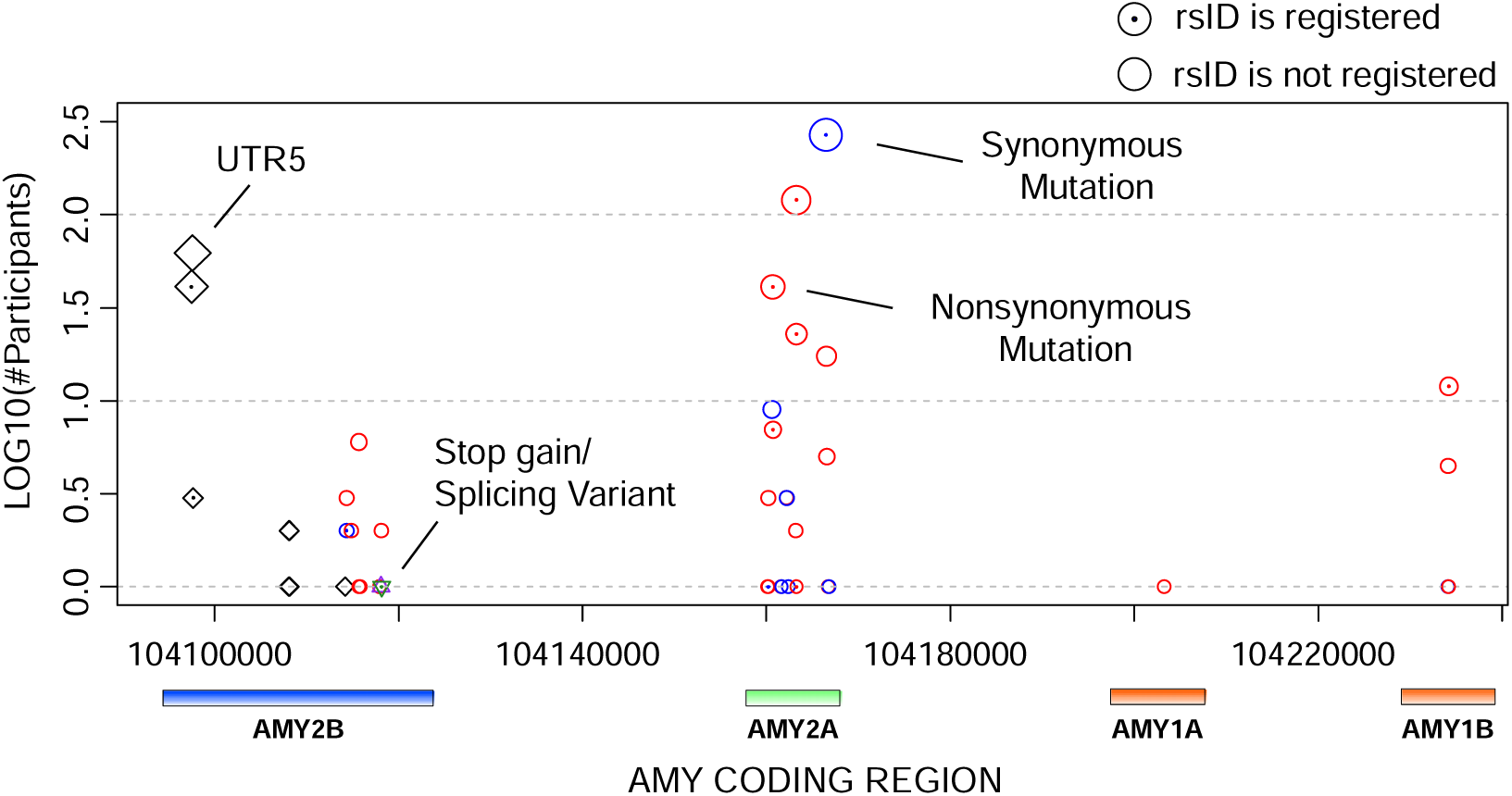
Distributions of SNPs on the exonic regions of AMY1, AMY2A, and AMY2B. In this figure, the vertical and horizontal axes indicate the number of participants with the corresponding SNPs and genomic positions including AMY1A, AMY2A, and AMY2B. Red, blue, green, black, and orange circles indicate nonsynonymous, synonymous, stop-gain (nonsense), splicing site, and 5′ prime untranslated region mutations, respectively. The dot at the center of the circles indicates that the SNPs have reference SNP IDs. Genes corresponding to the positions are illustrated at the bottom of the figure.

### Effect of AMY CN on HbA1c and BMI

Then, we performed a linear regression analysis of AMY1 and AMY2A CNs on HbA1c and BMI with the explained covariates. Therefore, AMY1-CNs and AMY2-CNs were positively associated with HbA1c and BMI, and their p-values for the first and second criteria of AMY1-CN and AMY2A-CN were 1.34E-2, 1.95E-2, and 2.76E-2 for HbA1c and 4.71E-1, 3.28E-2, and 6.25E-2 for BMI, respectively, as illustrated in Fig3. Using the predefined threshold (p = 0.05), we confirmed the statistically significant association of AMY1 and AMY2A CNs with HbA1c. However, only AMY1A in the second criterion tended to be associated with BMI. These results are partially consistent with those of a previous study (12); further, the association with HbA1c in the Japanese population seems to be a novel observation.

**Figure3.**
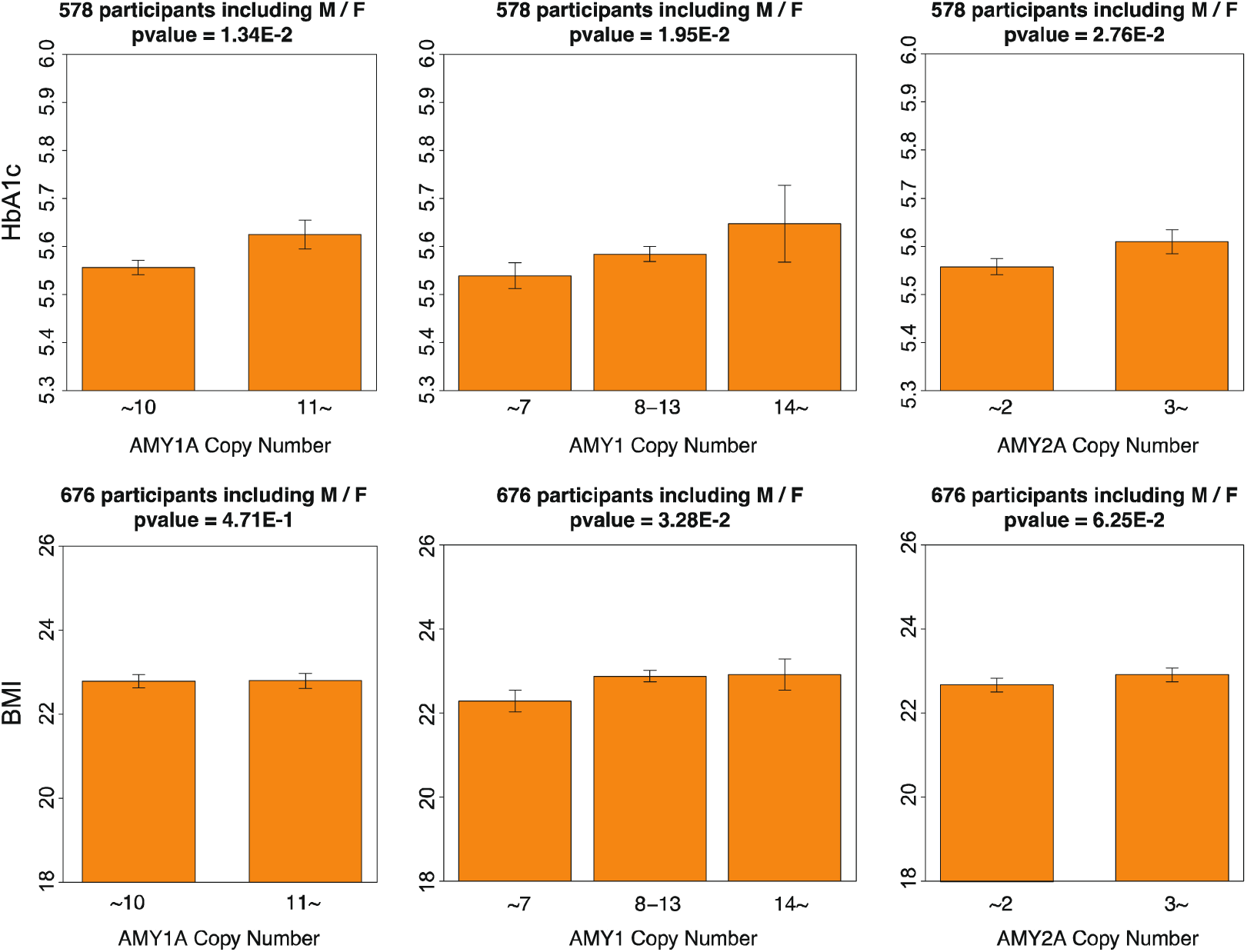
Association of HbA1c and BMI with AMY1A and AMY2A-CNs. In the upper and bottom parts of this figure, the associations of HbA1c and BMI with the criterion 1 and 2 of AMY1-CNs and AMY2-CNs are illustrated as barplots, respectively. The number of analyzed participants and p-values calculated using linear regression models with predefined covariates are displayed on the barplots.

### Association analysis between genus level microbiome composition and AMY CNs

In this study, we obtained the oral and gut microbiome composition of the participants in 2015, 2016, 2017, and 2018 through 16S rRNA sequencing. Among these, we extracted the data from participants whose WGS was also obtained to calculate AMY-CNs in 2014 or 2015. Considering the exclusion criteria, we selected 451, 487, 426, and 483 participants for the oral microbiome and 435, 475, 412, and 465 participants for gut microbiome analysis in 2015, 2016, 2017, and 2018, respectively. In the microbiome analysis, we excluded bacteria for which the sum of compositions of all participants was less than 1.0E-5 and (ii) those that were included in less than 3% of the participants each year. We thus retained 90 and 135 genus level bacteria for the oral and gut microbiome analyses, respectively. Thus, their predefined significant level with Bonferroni correction were 0.05 / 90 = 5.56E-4 and 0.05 / 135 = 3.70E-4, respectively. Because it was difficult to integrate microbiome composition data over the entire study period, we calculated integrated p-values using fixed-effect meta-analysis based on the estimates and standard errors. The obtained results are listed in Table 2; we extracted two prominent bacteria associated with AMY1-CNs, as illustrated in Fig.4. *Capnocytophaga* in the oral microbiome tended to be negatively associated with AMY1-CN (p = 1.38E-6). In addition, the *Dialister* composition in the gut microbiome increased and decreased with the increase in AMY1 CNs for males and females, respectively, and these differences were statistically significant (p = 1.02E-6 for males and 6.00E-6 for females). In particular, *Dialister* has been reported to be associated with carbohydrate metabolism and host diet; we could reproduce these results (18, 22). The top 10 bacteria and their p-values are presented in the supplementary material.

**Figure4.**
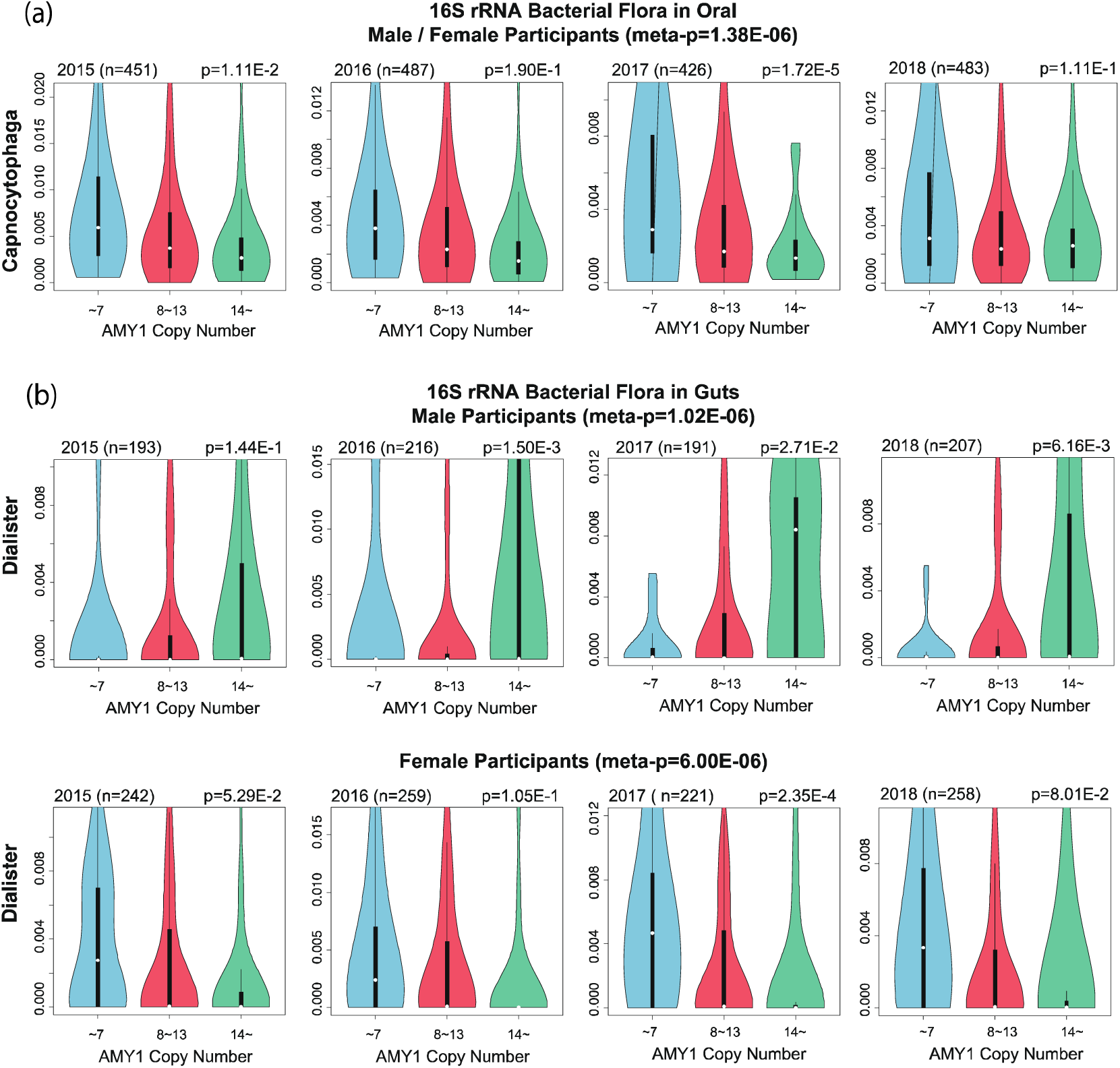
Association of the oral and gut microbiome with AMY1-CN. In the upper, middle, and bottom panels of this figure, associations of the composition of *Capnocytophaga* for males and females, *Dialister* for males, and *Dialister* for females are illustrated by violin-plot. The number of analyzed participants for each year and the p-values calculated using linear regression models with predefined covariates are displayed on the plots.

### Functional gene abundance analysis

Through the above experiments, AMY-CNs can be associated with HbA1c levels, BMI, and the oral and gut microbiomes. To further investigate the underlying mechanisms, we analyzed the abundance of functional gene sets in the oral and gut microbiome in association with AMY-CNs using shotgun microbiome sequencing. As we obtained data for 2015 and 2016, we considered participants whose WGS was obtained in 2014 and 2015. In this experiment, we generated the data by selecting the participants in 2015 and those participants in 2016 who did not participate in 2015, and integrating them into a single table. A total of 617 participants were thus included in this experiment. Because AMY-CNs determine the capacity of amylase secretion, which decomposes starch into dextrin and maltose, we focused on the starch degradation pathway. In the KEGG pathway database (23), we selected starch and sucrose metabolism (KEGG Identifier: map00500), amino sugar and nucleotide sugar metabolism (map00520), and Acarbose and validamycin biosynthesis (map00525) as potentially associated pathways. For this analysis, the total abundance of the gene set in each pathway was calculated for each participant, and a linear regression analysis with predefined covariates was also performed to analyze the association with AMY-CNs. Here, the predefined p-value with Bonferroni correction was 0.05 / 3 = 1.67E-2. As a result, the total abundance of genes in map0525 was negatively associated with AMY1-CN under the first and second criteria (p = 5.80E-4 and 3.97E-2, respectively) and with AMY2A-CN (p-value = 1.15E-3), as illustrated in Fig.5. Acarbose is an oral hypoglycemic drug used for treating type 2 diabetes, and this decrease could increase blood glucose levels.

**Figure 5.**
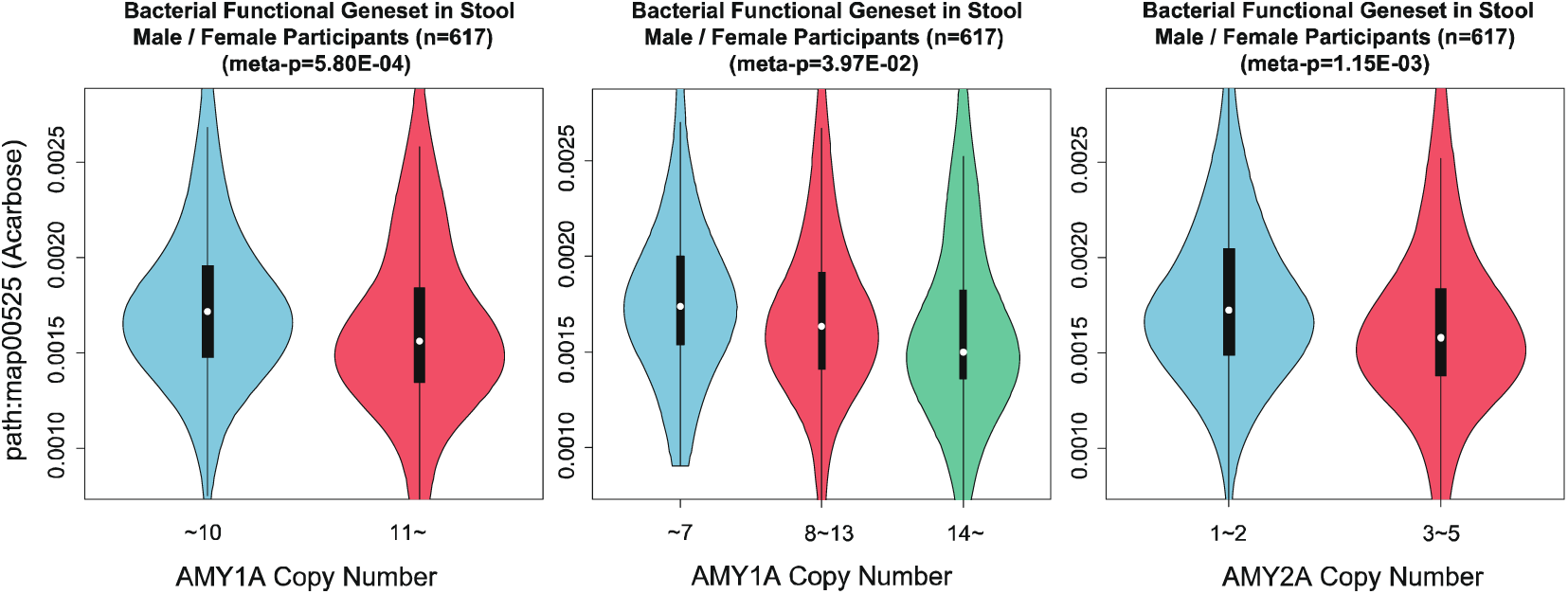
Associations of abundance of the Acarbose pathway gene set. The associations of the total abundance of the gene-set in the Acarbose pathway (KEGG: map00525) to criterion 1 and 2 of AMY1-CNs and AMY2-CNs are illustrated as violin-plots. The number of analyzed participants and p-values calculated by linear regression models with predefined covariates are displayed on the plots.

## Discussion and Conclusion

Several studies published in the last decade have shown that AMY1-CN is associated with obesity and diabetes. In contrast, several studies have reported that no significant association was observed among them. These conflicting results may have been influenced by the selection of populations and their lifestyles. For example, in people who eat a low starch diet, a high percentage of salivary amylase digests starch quickly and serum glucose levels become quickly and temporarily high at an early stage, whereas a low percentage of amylase can maintain a high serum glucose level. On the contrary, for people who eat large amounts of starch, a high percentage of salivary amylase may result in high glucose levels, whereas a low salivary amylase level may maintain moderate serum glucose levels. For most human beings, grains and rice are a major source of energy. Thus, it is important to elucidate the impact and mechanism of AMY-CNV on obesity and diabetes.

First, we assumed that the residents of Hirosaki City consume a relatively large amount of starch, even in Japan. According to a survey of rice consumption in two or more households by the Statistics Bureau, Ministry of Internal Affairs and Communications in Japan (https://www.stat.go.jp/data/kakei/5.html), the national average is 64.16 [kg/year], the consumption in Morioka city, which is close to Hirosaki city, is 71.61 [kg/year], whereas that in Tokyo is 53.16 [kg/year]; thus, the high starch consumption in this area might be related to this observation. Further, the AMY1A region has very few SNPs compared to the AMY2A and AMY2B regions. Thus, it seems that AMY1A exhibits diversity by copy number, and the AMY2A and AMY2B express diversity by mutation. However, in previous studies, more frequent SNPs in the AMY1 region have been reported; these results might thus be region-specific.

Similar to previous studies, we examined the association between AMY1-CN and HbA1c and BMI. The results showed that both HbA1c levels and BMI increased significantly with the increase in AMY1-CN. This is consistent with the finding of a positive correlation between the increase in AMY1-CN and BMI in the high starch intake group (12), but the association with HbA1c levels is a new finding in the Japanese population. Further, because the average AMY1-CN in this study was clearly greater than that in other studies, there could be a positive selection that favored more AMY1-CN in the past.

AMY-CN is reported to have a strong correlation with amylase secretion, and has a stronger effect on BMI and the oral and gut microbiome. As the amount of amylase secretion was not measured in this study, we examined the association between AMY1-CN and the oral and gut microbiome. Previously, the association of the human microbiome and AMY-CNs and the effect of microbiome translocation with obesity have been reported (16-18). Those studies have reported that *Prevotella* in the oral cavity and *Ruminococcus* in the gut increased with an increase in AMY1-CN. In our experiment, although not statistically significant, similar tendencies were observed. In particular, in this study, we found that *Capnocytophaga* decreased significantly in the oral cavity as AMY1-CN increased. *Capnocytophaga* is known to be associated with periodontal disease and could be affected by the amount of starch decomposition; however, to the best of our knowledge, no study has been published regarding its associations. In addition, *Dialister* increased significantly in men and decreased significantly in women with an increase in AMY1-CN. *Dialister* has been reported to

increase in obese women, but we show a strong correlation with AMY1-CN rather than with BMI and HbA1c levels. The amount of *Dialister* may be thus directly affected by the amount of amylase secreted, which could result in obesity and other consequences.

Finally, we tried to elucidate the mechanism underlying the effect of the oral and gut microbiome on obesity and diabetes from the viewpoint of changes in the abundance of pathway-based functional genesets. The results showed that the total amount of acarbose and validamycin biosynthesis (map00525) decreased significantly with an increase in both AMY1-CN and AMY2A-CN. Acarbose is an oral hypoglycemic drug used for the treatment of type 2 diabetes, which delays starch breakdown to glucose by inhibiting the action of α-glucosidase. Thus, glucose is absorbed more slowly into the body, and postprandial hyperglycemia can be suppressed. In other words, in the group with a small number of AMY-CN, the expression of acarbose in the gut can be increased and the increase in blood glucose is suppressed; in contrast, with a large number of AMY-CN, the expression of acarbose is decreased and the increase in blood glucose is promoted. Through this mechanism, we could partially explain the causes of an increase in HbA1c levels and BMI in the group with a large AMY-CN.

Within this study, we considered a particularly limited population in Japan, and so, these results may be specific to lifestyle, genome, and the oral and gut microbiome, and may not be applicable to other populations. However, even in such a specific population, our findings are partially consistent with previous research, and novel findings on AMY-CN have been further obtained.

## Materials and Methods

### Ethics statement

This study design was approved by the Hirosaki University Hospital Institutional Review Board (Protocol Number Num. 2017-026). Written informed consent was obtained from all the patients in this study. No animal experiments were conducted in this study.

### Sample collection and preparation

We analyzed the data obtained from the Hirosaki Center of Innovation (COI) health promotion project, which has collected cohort data from the year 2005 in Hirosaki (the population is approximately 10, 000) in northern Japan. Each year, approximately 1,000 people have their health status checked through a free participation program. The dataset actually has numerous missing data because the participants do not necessarily undergo a checkup each year. Thus, the number of analyzed participants was displayed in each experiment. Questionnaires and blood tests were collected over the entire study period, and single nucleotide polymorphisms (SNPs) were identified for participants in 2014, 2015, 2016, and 2017; WGS analysis was performed for participants in 2014 and 2015; 16S rRNA sequencing of the oral and gut microbiomes was performed for participants in 2015, 2016, 2017, and 2018, and whole-genome shotgun sequencing of the oral and gut microbiomes was performed for participants in 2015 and 2016.

### Participant selection and covariate assessment

The main exclusion criteria were as follows: (1) participants were younger than 18 years of age or were 55 older; (2) those with an estimated glomerular filtration rate (eGFR) below 60 [mL/min/1.73 m^2^] as calculated from serum creatinine levels and age at admission; (3) those with serum total bilirubin levels above 2 [mg/dl], (4) those with serum albumin levels above 35 [g/dl], (5) those with a BMI above 35 or below 18, (6) those who were prescribed medicine for diabetes mellitus as per their questionnaires, (7) those who were prescribed antidiabetic or antibiotic medication as per their questionnaires, for the analysis of oral and gut microbiome; and (8) those with missing information regarding any of their covariates.

The participant covariate information was obtained from the blood value tests and questionnaires. The questionnaires were administered upon induction into the study. (a) Age and (b) sex information were obtained from questionnaires (sex was also checked using genetic data), (c) smoking status was classified as never (0) and current and former (1); (d) drinking status was classified as non-drinker (0), and current and former drinker (1). Participants who used medication for (e) high blood pressure, (f) dyslipidemia, (g) steroids, and (h) antibiotics were flagged as non-use (0) and use (1). Finally, we performed principal component analysis (PCA) for genotyping using the algorithm implemented in smartpca (24), and used the top five principal components (PCs) as covariates to represent the population structure based on the genetic correlations among individuals.

### SNPs and whole genome sequencing

DNA was purified from peripheral whole blood using a DNA extraction kit (QIAamp 96 DNA Blood Kit (QIAGEN, Hilden, Germany)) and was extracted from plasma pellets for SNP arrays and whole genome sequencing, respectively. SNP information was obtained from Japonica Array (Toshiba, Tokyo, Japan) (25) and comprised population-specific SNP markers designed from the 1070 whole genome reference panel (26), and whole genome sequencing was performed by Takara Bio Corporation (Shiga, Japan).

### Analysis of AMY1, AMY2A, and AMY2B copy number variations

To detect AMY1, AMY2A, and AMY2B CNVs from the WGS data, we used AMYCNE (21), which was developed to estimate the AMY CNV from sequencing data. The input data for this software was prepared using TIDIT (20) which was also developed to clarify the structural variants.

### SNP analysis for AMY1, AMY2A, and AMY2B regions

For SNP detection, we applied the Genomon pipeline (https://github.com/Genomon-Project), which has been developed to investigate mutations from exome/whole genome sequencing data. The filter conditions were as follows: minor allele frequency of 0.02 or higher, minimum read depth higher than 8, and 10% posterior percentile of the minor allele count based on a binomial distribution greater than 0.02, and a minimum base quality of 15 or higher.

### Microbiome assay

Microbiome samples were obtained from participants’ tongue plaques and stool samples for oral and gut microbiomes, respectively, and were stored at -80°C until use. The detailed library preparation method, including PCR conditions, has been described in a previous paper (27). Briefly, the samples were mixed with zirconia beads and lysed using a FastPrep FP100A instrument (MP Biomedicals, Santa Ana, California, USA). DNA was extracted from the bead-treated suspensions using a Magtration System 12GC and GC series MagDEA DNA 200 (Precision System Science, Chiba, Japan) and amplified. The 16S rRNA gene amplicons covering the V3–V4 region were sequenced using Pro341F and Pro805R primers. Sequencing was performed using a paired-end, 2×250-base pair cycle run on an Illumina MiSeq sequencing system. Shotgun whole genome sequencing was performed on the HiSeq2500 instrument (Illumina, San Diego, California, USA) with 101□bp paired-end reads.

### Calculation of genus level microbiome abundance profiles from 16S rRNA sequences

Adapter sequences and low-quality bases at the 3′-end (threshold = 30) were trimmed using Cutadapt (version: 1.13) (28) from paired-end reads obtained using the Illumina MiSeq sequencing system. In addition, the reads containing N bases and reads shorter than 150 bases were removed. The remaining reads were merged into single reads, and those with a length of 370 to 470 bases were removed using the VSEARCH (29) fastq_mergepairs subcommand (version: version 2.4.3). After merging, we removed reads that were expected to have one or more base errors from the base quality. After removing chimeric reads using the VSEARCH (29) uchime_denovo subcommand, the reads were clustered with 97% sequence identity and the taxa of each cluster were identified using the RDP Classifier (git commit hash: (commit hash: 701e229dde7cbe53d4261301e23459d91615999d)) (30). At this time, the result with a confidence value < 0.8, was treated as unclassified. The relative abundance of each taxon was then calculated by dividing the number of reads belonging to the cluster to which the taxon was assigned by the total number of reads.

### Calculation of functional gene abundance from shotgun metagenomic sequencing

Adapter sequence and low-quality bases at the 3’-end (threshold = 30) were trimmed using Cutadapt (version: 1.13) from 2×100-base paired-end reads obtained using the Illumina HiSeq 2500 system. Reads containing N bases and reads shorter than 80 bp were removed. Reads from the host genome were also removed using BWA-MEM (version 0.7.15) (31) with GRCh38. From the remaining reads (referred to as non-host reads), contigs were generated by metagenomic assembly using MEGAHIT (version 1.1.1) (32). The coding regions of genes on these contigs were predicted using Prodigal (version 2.6.3) (33). After mapping the non-host reads to contigs using BWA-MEM, the relative abundances of the genes were calculated from the average depth of the gene region. KEGG pathway annotations were mapped to genes using a sequence similarity search with MMseqs2 (version: 1d2579627f43662ecaaa0778bd348fc35048976a) (34). We used 1E-10 as the E-value threshold for the similarity search.

### Statistical Analysis

In the first criterion for AMY1-CN, participants were divided into low (AMY1-CN range 3–10) and high (AMY1-CN range 11–24) groups, which were separated at the center line. In the second criterion for AMY1-CN, participants were divided into the low (AMY1-CN range, 3–7), medium (range 8-13), and high (range, 14–24) groups. The minimum copy number above 10 percentile and the maximum copy number below 90 percentile of AMY1-CN distribution was 7 and 14, respectively. Similarly, for analyzing AMY2A-CN, participants were divided into low (AMY2A-CN range 1–2) and high (AMY2A-CN range 3–5) groups at the center line. We could not analyze the effect of AMY2B-CN because almost all patients had two AMY2B-CNs. For these groups, linear regression analysis using the above-explained covariates was applied to detect statistical significance. A p-value less than 0.05 was considered to indicate statistical significance when considering the corrections for multiple tests.

## Data Availability

The datasets supporting the current study have not been deposited in a public repository [because the data are not public] but are available from the corresponding author, S.I., on request.

## Acknowledgments

This work was supported by JST COI Grant Number JPMJCE1302. This research used the supercomputing resources provided by the Human Genome Center, Institute of Medical Science, University of Tokyo (http://sc.hgc.jp/shirokane.html).

## Authors’ contributions

T.H. designed the study, analyzed the data, interpreted the data, and wrote the manuscript. M. K., R.Y., and N.S. analyzed the data, interpreted the data, helped write the manuscript, and commented on the drafts. M.A., J.S., I.T., and K.M. performed the medical checkups and gathered and summarized the data. S.I. interpreted the data, helped write the manuscript, commented on the drafts, and supervised the study. All authors have read and approved the final manuscript.

## Supplementary Material

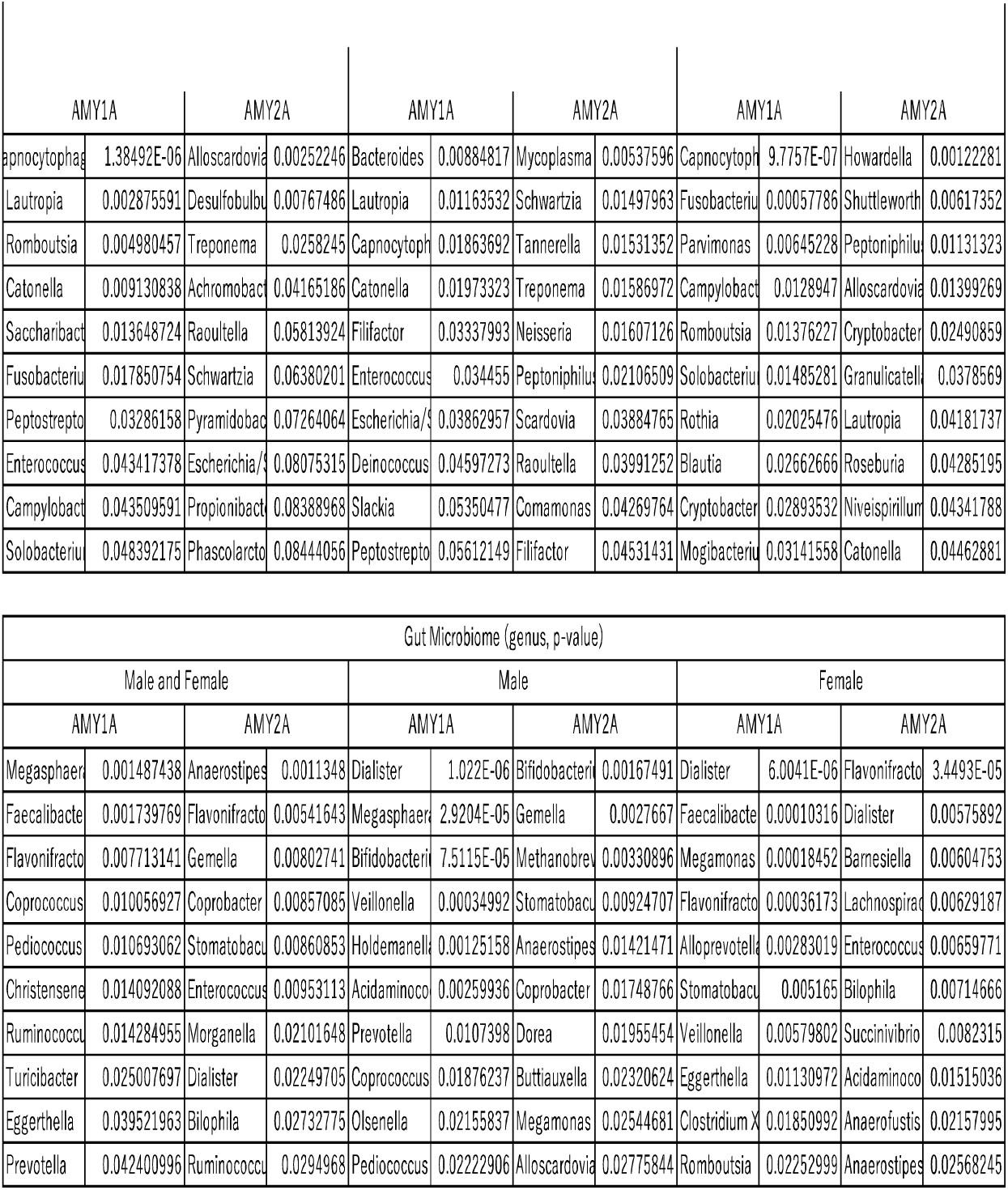

