## Supplemental Table for "Impact of Salivary and Pancreatic Amylase Gene Copy Numbers on Diabetes, Obesity, and Functional Profiles of Microbiome in Northern Japanese Population"

| Oral Microbiome (genus, p-value) | | | | | | | | | | | |
|---|---|---|---|---|---|---|---|---|---|---|---|
| Male and Female | | | | Male | | | | Female | | | |
| AMY1A | | AMY2A | | AMY1A | | AMY2A | | AMY1A | | AMY2A | |
| apnocytophag | 1.38492E-06 | Alloscardovia | 0.00252246 | Bacteroides | 0.00884817 | Mycoplasma | 0.00537596 | Capnocytoph | 9.7757E-07 | Howardella | 0.00122281 |
| Lautropia | 0.002875591 | Desulfobulbu | 0.00767486 | Lautropia | 0.01163532 | Schwartzia | 0.01497963 | Fusobacteriu | 0.00057786 | Shuttleworth | 0.00617352 |
| Romboutsia | 0.004980457 | Treponema | 0.0258245 | Capnocytoph | 0.01863692 | Tannerella | 0.01531352 | Parvimonas | 0.00645228 | Peptoniphilus | 0.01131323 |
| Catonella | 0.009130838 | Achromobact | 0.04165186 | Catonella | 0.01973323 | Treponema | 0.01586972 | Campylobact | 0.0128947 | Alloscardovia | 0.01399269 |
| Saccharibact | 0.013648724 | Raoultella | 0.05813924 | Filifactor | 0.03337993 | Neisseria | 0.01607126 | Romboutsia | 0.01376227 | Cryptobacter | 0.02490859 |
| Fusobacteriu | 0.017850754 | Schwartzia | 0.06380201 | Enterococcus | 0.034455 | Peptoniphilus | 0.02106509 | Solobacteriu | 0.01485281 | Granulicatella | 0.0378569 |
| Peptostrepto | 0.03286158 | Pyramidobac | 0.07264064 | Escherichia/S | 0.03862957 | Scardovia | 0.03884765 | Rothia | 0.02025476 | Lautropia | 0.04181737 |
| Enterococcus | 0.043417378 | Escherichia/S | 0.08075315 | Deinococcus | 0.04597273 | Raoultella | 0.03991252 | Blautia | 0.02662666 | Roseburia | 0.04285195 |
| Campylobact | 0.043509591 | Propionibacte | 0.08388968 | Slackia | 0.05350477 | Comamonas | 0.04269764 | Cryptobacter | 0.02893532 | Niveispirillum | 0.04341788 |
| Solobacteriu | 0.048392175 | Phascolarcto | 0.08444056 | Peptostrepto | 0.05612149 | Filifactor | 0.04531431 | Mogibacteriu | 0.03141558 | Catonella | 0.04462881 |

| Gut Microbiome (genus, p-value) | | | | | | | | | | | |
|---|---|---|---|---|---|---|---|---|---|---|---|
| Male and Female | | | | Male | | | | Female | | | |
| AMY1A | | AMY2A | | AMY1A | | AMY2A | | AMY1A | | AMY2A | |
| Megasphaera | 0.001487438 | Anaerostipes | 0.0011348 | Dialister | 1.022E-06 | Bifidobacteriu | 0.00167491 | Dialister | 6.0041E-06 | Flavonifracto | 3.4493E-05 |
| Faecalibacte | 0.001739769 | Flavonifracto | 0.00541643 | Megasphaera | 2.9204E-05 | Gemella | 0.0027667 | Faecalibacte | 0.00010316 | Dialister | 0.00575892 |
| Flavonifracto | 0.007713141 | Gemella | 0.00802741 | Bifidobacteriu | 7.5115E-05 | Methanobrev | 0.00330896 | Megamonas | 0.00018452 | Barnesiella | 0.00604753 |
| Coprococcus | 0.010056927 | Coprobacter | 0.00857085 | Veillonella | 0.00034992 | Stomatobacu | 0.00924707 | Flavonifracto | 0.00036173 | Lachnospirac | 0.00629187 |
| Pediococcus | 0.010693062 | Stomatobacu | 0.00860853 | Holdemanella | 0.00125158 | Anaerostipes | 0.01421471 | Alloprevotella | 0.00283019 | Enterococcus | 0.00659771 |
| Christensene | 0.014092088 | Enterococcus | 0.00953113 | Acidaminoco | 0.00259936 | Coprobacter | 0.01748766 | Stomatobacu | 0.005165 | Bilophila | 0.00714666 |
| Ruminococcu | 0.014284955 | Morganella | 0.02101648 | Prevotella | 0.0107398 | Dorea | 0.01955454 | Veillonella | 0.00579802 | Succinivibrio | 0.0082315 |
| Turicibacter | 0.025007697 | Dialister | 0.02249705 | Coprococcus | 0.01876237 | Buttiauxella | 0.02320624 | Eggerthella | 0.01130972 | Acidaminococ | 0.01515036 |
| Eggerthella | 0.039521963 | Bilophila | 0.02732775 | Olsenella | 0.02155837 | Megamonas | 0.02544681 | Clostridium X | 0.01850992 | Anaerofustis | 0.02157995 |
| Prevotella | 0.042400996 | Ruminococcu | 0.0294968 | Pediococcus | 0.02222906 | Alloscardovia | 0.02775844 | Romboutsia | 0.02252999 | Anaerostipes | 0.02568245 |
